# *PABPN1* loss-of-function in oculopharyngeal muscular dystrophy primarily impacts APA-shift in muscle transcripts

**DOI:** 10.1101/2023.08.17.23294024

**Authors:** Milad Shademan, Hailiang Mei, Yavuz Ariyurek, Susan Kloet, Vered Raz

## Abstract

Alternative polyadenylation (APA) at the 3’UTR of transcripts contributes to the cell transcriptome. APA is suppressed by the nuclear RNA binding protein, PABPN1. Aging-associated reduced PABPN1 levels in skeletal muscles lead to muscle wasting. Muscle weakness in oculopharyngeal muscular dystrophy (OPMD) is caused by short alanine expansion in PABPN1 exon1. The expanded PABPN1 forms nuclear aggregates, an OPMD hallmark. Whether the expanded PABPN1 affect APA and how contributes to muscle pathology is unresolved.

To investigate these questions, we developed a procedure including RNA library preparation and a simple pipeline calculating ‘APA-shift’ ratio as a readout for PABPN1 function. Using the mouse OPMD model we demonstrate similar results between previously published PAS utilization and ‘APA-shift’ results. Studying APA-shift in two OPMD models and in OPMD patients we show that the expression of the expanded PABPN1 does not correlate with APA-shift. Instead, APA-shift is correlated with reduced expression levels of PABPN1 isoforms, amongst the isoform lacking exon1. Further we show that with our protocol APA-shift is enriched in muscle transcripts, moreover in OPMD patients. We suggest that muscle weakness in OPMD is caused by *PABPN1* loss-of-function leading to APA-shift that primarily affects in muscle transcripts.

## Introduction

The landscape of mRNAs in eukaryote cells is highly impacted by the 3’ untranslated regions (3’UTR) of transcripts. The length of the 3’UTR affects multiple layers of mRNAs processing including nuclear export, translation efficiency, and stability or decay [1, 31]. The 3’UTR length is determined by cleavage of the polyadenylation site (PAS), a long 3’UTR is generated from a distal PAS, whereas utilization of proximal PAS generates the short isoform. Around 70% of genes have two or more PASs at the 3’UTR [12], PASs can be discriminated between canonical and alternative PASs (APAs), from the later alternative transcripts are formed [12, 25]. In general, longer 3’UTR transcripts have higher translation efficiency [37], whereas short 3’UTR have reduced abundance in the heavy polysomes and reduced translation efficiency [24]. APA generates alternative transcripts that augment the cell transcriptome and protein landscape.

APA is regulated by RNA processing and RNA binding proteins [15, 39], some are ubiquitously expressed and others have a cell or tissue-specific expression [26, 34]. Consequently, alternative transcripts generated from APA can show a cell-type specific pattern and can impact biological processes in health and disease [7, 39].

Among the RNA binding proteins regulating APA site utilization, the PolyAdenosine (PolyA) RNA Binding Protein Nuclear 1 (PABPN1) suppresses the APA. PABPN1 depletion causes genome-wide APA site utilization and shorter transcripts [12, 19]. These shorter transcripts have a lower abundance in polyribosomes, impacting translation efficiency [24].

PABPN1 is a vital protein and is ubiquitously expressed in all cells. PABPN1 levels are reduced during normal aging in skeletal muscles [2]. Reduced PABPN1 levels were also reported in oculopharyngeal muscular dystrophy (OPMD), an adult-onset autosomal dominant myopathy. OPMD is caused by a trinucleotide alanine expansion mutation (polyAla) in the first exon of the gene encoding PABPN1 [6]. The expanded PABPN1 forms nuclear aggregates and entrap other nuclear proteins and mRNAs, a histopathological mark of OPMD [38]. A mouse model overexpressing the expanded PABPN1 in the skeletal muscles exhibits muscle wasting [40], concomitant with genome-wide APA alterations [12, 19]. This mouse model was instrumental to demonstrate that PABPN1 aggregation leads to muscle pathology concurrently with APA. So far, genome-wide APA has not been reported in OPMD patients, and it is unclear to what extent PABPN1 molecular function contributes to OPMD pathology.

To address this question, we developed a wet lab and computational procedure reporting changes in 3’UTR length of mRNA isoforms, named ‘APA-shift’. We show that this method is highly robust and specifically beneficial to detect APA-shift in muscle transcripts. Applying the method to OPMD models and to RNA from OPMD patients we demonstrate that APA-shift correlates with reduced levels of PABPN1 isoform lacking the polyAla expansion rather than by the expression of mutant PABPN1. Our study

## Materials & Methods

### Mouse

Tibialis anterior muscles from A17.1 and FVB males 18 months old were collected, stored and bulk RNA was extracted as previously reported [40].

### Mouse muscle cells

stable cells expressing Ala10 or Ala17 PABPN1 under muscle actin promoter were reported [29]. Bulk RNA was extracted from fused cultures as detailed in [29].

### Human

vastus lateralis muscle was collected from controls and OPMD patients, genetically confirmed, previously detailed [13]. For the APA-seq, 12 controls and 10 OPMD were selected. Four controls were excluded due to poor sequencing quality. Age and gender of OPMD and controls are listed in Table S1.

All RNAs were stored at −80 and RNA integrity was determined prior to cDNA library preparation. The samples and models that were used for RNA sequencing and down-stream analysis are listed in Table S2A.

### Library preparation and RNA sequencing

The generation of cDNA for the RNA sequencing data was carried out with the mcSCRB-seq protocol [3], with the following modifications to the workflow: instead of single cells, we used 1-100 ng isolated total RNA, 7.5% PEG8000 was replaced with 0.5% Polyvinylpyrrolidone (PVP) and the oligodT primer was replaced with the SCIFI_LIG384_001 primer (Table S2C, also described in [11]).

All samples are barcoded during reverse transcription using a well barcode (Table S2C). After reverse transcription, the full-length cDNA was either unamplified (1C) or underwent 7 cycles of amplification (7C) using 2x Kapa HiFi HotStart mix (Roche). cDNA was then pooled, and sequencing libraries were generated using the Kapa Hyper Plus kit (Roche) using a standard protocol. Because the 1C libraries were not pre amplified we added more cycles, namely 14 cycles and the p7 primer was replaced with i7 Illumina barcode per pool. After an additional double SPRI size selection, the Illumina library was quantified using Qubit and checked on an Agilent Labonachip for the size distribution. Illumina paired-end sequencing of the library was done on a NovaSeq 6000 using paired-end 150bp sequencing and v1.5 chemistry. After sequencing Read 1 will contain the cDNA information and read 2 will contain only the UMI.

For the pipeline to run effortlessly the UMI in the read2 fastq file was added to the beginning of the same read id in the read1 fastq file. The modified Read1 fastq file starts with the UMI and is followed by the cDNA sequence, which was used in the RNA-seq pipeline. For the demultiplexing the pooled samples we used the well barcode from the oligodT primer using Ultraplex (https://github.com/ulelab/ultraplex) [42]. A summary of QC analysis of all samples is found in Table S2B.

### RNAseq analysis and APA-shift calculation

To process the APA-seq data to establish the APA-shift profiles in both our human and mouse studies, we established a data preprocessing pipeline (as depicted in Figure. S1). All mouse and human reads in FASTQ format are first filtered using Cutadapt (v2.10) to remove all remaining adapter sequences. Then the reads were aligned to the Ensembl transcriptome version 104 using STAR (v2.7.5a) including UMI based deduplication using UMI-Tools (v1.1.1) to generate transcriptome-based alignment in BAM format so that we could visualize using IGV. By inspecting these alignment files in 3’ UTR using IGV, we could observe clear enriched signals indicating both proximal and distal APA sites (Figure 1A). Confirmed by the visual inspection in IGV, we used the same Ensembl gene annotation version 104 to create a customized 3’ UTR annotation GTF file for all Ensembl transcripts with annotated 3’ UTR. In this GTF file, 3’ UTRs are split in the middle to create a proximal and a distal region. With this APA proximal and distal annotation file and the human and mouse transcriptome-based alignment file as input, we quantified the APA enrichment signal at both proximal and distal regions using featureCounts (v2.0.1) with option “-M” to include multiple mapped reads.

**Figure 1.**
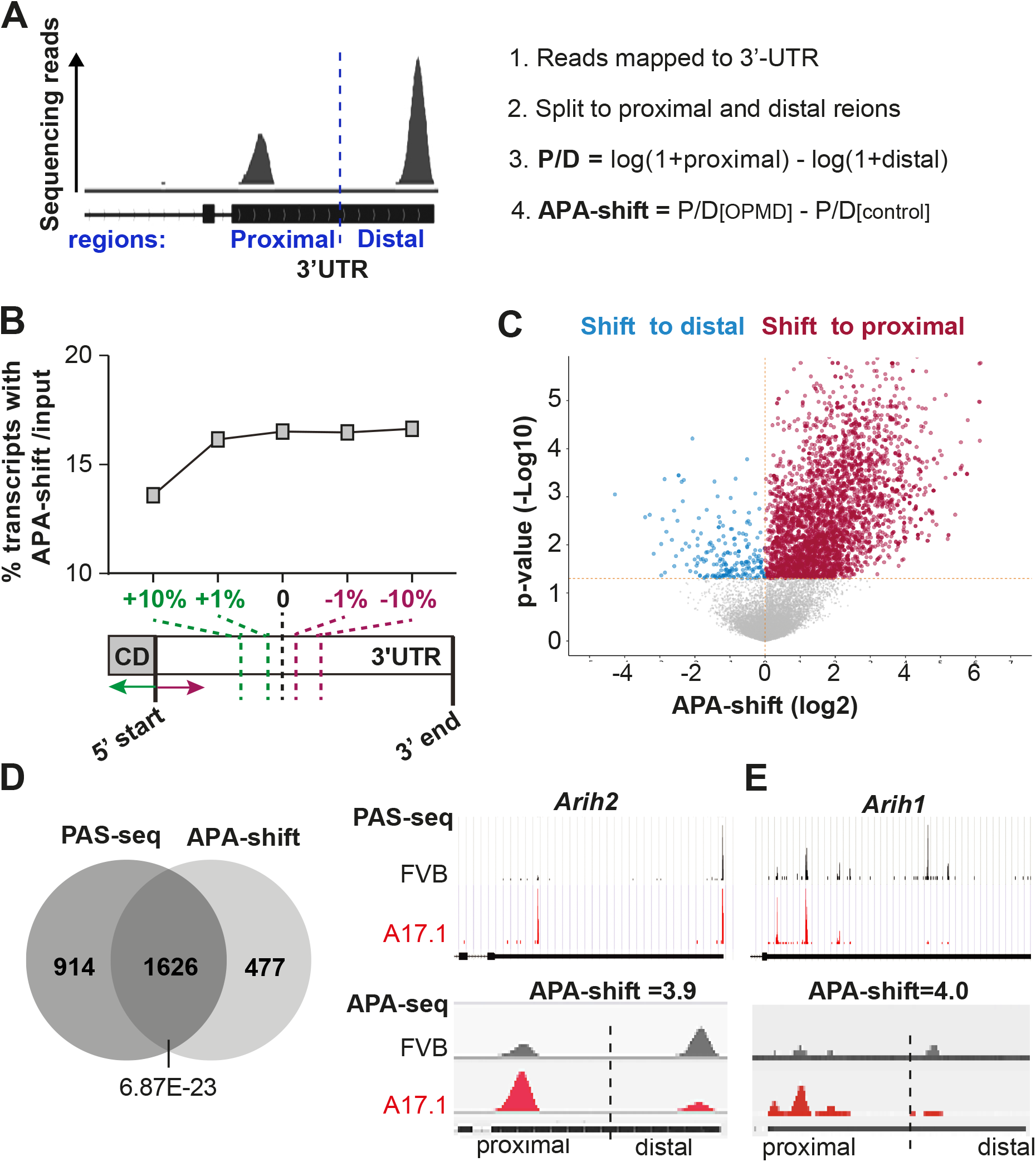
APA-shift calculation. **A.** Left: IGV from a 3’UTR transcript example for the APA-seq. The split point between proximal and distal regions is denoted with a dashed line. Right: a summary of major steps in the APA-shift pipeline calculation. **B.** Shortening and extending split point 3’UTR regions effect on APA-shift. A schematic presentation of 3’UTR splitting into proximal and distal regions: in the middle (depicted by dashed black line), or extension to towards 5’ sequences (in green) or shortening towards 3’sequences (in red) by 1% and 10% relative to the middle point, is depicted under the dot plot. Dot plot shows the percentage of transcripts with APA-shift **C.** Volcano plot of APA-shift in A17.1 vs. FVB mouse using a generic protocol. Mean -log fold change and *P-*values are from N=5 per genotype. Transcripts with a switch to proximal are highlighted in red and a switch to distal in blue. **D.** Integrative genomic viewer plots of the sum of reads at the 3’UTR for two example significant genes *Arih2* and *Arih1* 3’UTR from polyadenylation site (PAS) sequencing (de Klerk 2012) and APA seq in FVB and A17.1. The 3’UTR is depicted by the black horizontal bar, and the vertical dashed line indicates the 0 position. The average APA-shift fold change (AVR FC) for *Arih1* ENSMUST00000171975 and *Arih2* ENSMUST00000013338 are depicted. **E.** Venn diagram shows the overlap of transcripts with significant APA-shift between PAS and APA sequencing. *P-*value for overlap significance was determined with chi-square test.

Transcripts were excluded according to the exclusion criteria in Table S2. Raw counts were used to calculate the ratio between proximal and distal (P/D) in R, using the following equation: log (1+P) – log (1+D). APA-shift, in a log scale, was calculated between OPMD and control as P/D [OPMD]-P/D[control]. A ratio larger than 0 indicates a shift to proximal and smaller than 0 a shift to distal. A significant shift was considered by statistical analysis. The power of the statistical test varied was adjusted to the number of samples and the expected variation.

For human data, we incorporated age and gender as covariates in a model matrix. The P/D ratios were corrected on data corrected for age and gender using a linear model in ‘limma’, and APA-shift was calculated on the corrected data. A list of transcripts with APA-shift in mouse, cell, and human samples is found in Table S3).

To assess if splitting the 3’UTR in the middle is a robust choice, we examined APA-shift results by extending or shortening the annotated 3’UTR. Four different shortening or extension factors were examined. A shortening refers to moving the starting position of 3’UTR towards the 3’ end, while extension refer to moving the starting position of 3’UTR towards the 5’ end. The 3’ end-position of a transcript was unchanged. The resulted shortened or extended 3’UTR were split half-half, defining the proximal or distal region. Per direction, 10% and 1% moving factors were analyzed. Counts normalized to the mean were used to calculate expression levels.

### Reverse transcription quantitative polymerase chain reaction (RT-qPCR)

RT-qPCR was conducted on RNA extracted from the control and OPMD muscles. For cDNA synthesis, 500 ng RNA was reverse transcribed using the QuantiTect Reverse Transcription Kit (QIAGEN) and random primers, following the manufacturer’s instructions. Subsequently, qPCR amplification was performed with QuantiNova SYBR Green kit (QIAGEN) using 5 ng RNA, with technical duplicates, utilizing a standard amplification protocol at a melting temperature (TM) of 60°C. Samples with CT values above 35 were excluded from the analysis to eliminate potential noise. The average CT values from the technical duplicates was used for ddCT calculation. PABPN1 Exon-4-5 (ENSG00000100836) was normalized to HPRT, and isoforms 201 (ENST00000216727), 202 (ENST00000397276) and 207 (ENST00000556821) to exon 3-4. Isoforms 001 and 004 were targeted with specific primer sets, designed to cover different exons, thereby amplifying only mRNA molecules. Primer sets were designed with NCBI Primer design tool (https://www.ncbi.nlm.nih.gov/tools/primer-blast/) and primers sequence is in Supplementary Table S2D.

### Other resources

PAS-seq from A17.1 (5+5) and OPMD (4 patients and 4 controls) muscles was generated as detailed in [12]. Bam files of PAS-seq were used to visualize the genomic localization of reads peak. Transcripts with APA site utilization in A17.1 are from Table S2 [12].

### Statistical and enrichment analyses

Statistical analyses were conducted in R 4.2.3, and GraphPad Prism 9.3.1. Statistical significance of APA-shift was assessed by applying the Benjamini-Hochberg (BH) method to adjust the *P-*values for controlling the FDR.

The list of transcripts encoding for muscle proteins (named here muscle transcripts, Table S4) was downloaded from AmiGO2 geneontology.org using the Gene Ontology database (<u>AmiGO 2 - GO Wiki</u> (geneontology.org). The GSEA [36] V.4.3.2 was used for gene set enrichment analysis and DAVID [17] V.2023q1, for Gene ontology analysis considering the Bonferroni-corrected *P-*value in human and A17.1 mice.

Visualization of reads at the 3’UTR was carried out with the Integrative Genomics Viewer (IGV) 2.16.0.

## Results

### APA-shift calculation captures real-time APA dynamics in muscle genes

Aiming to detect APA-shift in OPMD conditions, we developed a computational pipeline that quantifies global changes in PAS utilization at the 3’UTR of transcripts from a standard poly(A) library preparation of the 3’UTR. We generated libraries from the A17.1 mouse model, that overexpresses the expanded PABPN1 in skeletal muscles, and APA was previously demonstrated by others and us [12, 19] RNA sequencing from these libraries was used to develop and optimize our pipeline. Since long transcripts are generated from the distal PAS and short transcripts from a proximal PAS at the 3’UTR, we defined proximal and distal regions based on a split in the middle of the 3’UTR and calculated the ratio between sum of reads from proximal and distal regions (Fig. 1A). The ratio between the proximal to the distal regions (P/D, Fig. 1A) and the ratio between OPMD conditions and control resulted in a value we named as ‘APA-shift’. Since multiple transcripts could belong to proximal and/or distal regions, and a split in the middle is an arbitrary choice, we assessed if moving the 3’UTR start point towards 5’ of the transcript (named as extending) or towards 3’ of the transcript (named as shortening) will impact global APA-shift results (Fig. 1B). For the shortening by 1% or 10%, and the extension by 1%, after APA-shift recalculation in all 3’-UTR transcripts, the percentage of transcripts with APA-shift was unchanged (Fig. 1B). In contrast, the extension of the 3’UTR by 10% included longer non-3’UTR sequences in the proximal region, resulted in reduced proportion of transcripts with APA-shift (Fig. 1B). This is expected as our library preparation was focused on short sequences starting from the poly(A) tail. This analysis shows that shifting the 3’UTR split point around the middle of the 3’UTR did not impact the percentage of transcripts with APA-shift on a transcriptome scale. Therefore, we continued with a split of the middle of the annotated 3’UTR.

In the A17.1 most transcripts showed a shift to proximal (Fig. 1C), agreeing with enhanced proximal APA in A17.1 [12, 19]. In a previous study we employed PAS-seq to determine APA site utilization in A17.1 [12]. The overlap between transcripts with PAS-seq and transcripts with APA-shift in A17.1 was highly significant, 77.3% of the APA-shift transcripts overlapped with PAS-seq (Fig. 1D). We then also compared reads signal at the proximal and distal 3’UTR regions in our study to the PAS-seq. We selected two transcripts for which PAS-seq was validated [12, 28]: *Arih2* (ENSMUST00000013338) has only two PAS at the 3’UTR, and *Arih1* (ENSMUST00000171975) has multiple PAS at the 3’UTR (Fig. 1E). For both the localization of signal reads at proximal and distal regions was highly similar between the two procedures (Fig. 1E). Thus, APA-shift can reproduce results obtained by APA-seq. This confirms that our simple APA-shift protocol is valid to report changes in PAS utilization. We continued with APA-shift analysis using the option where the 3’UTR is divided in the middle into the proximal and distal regions.

To capture real-time changes in APA, we optimized the library preparation protocol and compared the standard protocol using 7 amplification cycles (named here as ‘7C’) to a protocol employing 1 amplification cycle (named here ‘1C’). As expected, the percentage of mapped reads was higher with the 7C protocol, however, the percentage of reads after UMI-based deduplication was higher with the 1C protocol (Table S2B; 64% ±8.4, and 84% ±4.1, respectively). The 1C protocol resulted in a library depth ∼2-folds smaller than the 7C, but the average transcript size was 13% larger (Fig. 2A). A principal component analysis (PCA) showed similar differences between A17.1 and FVB samples (Fig. 2SA) in 7C and 1C protocols. Yet, with the 7C protocol, the average transcript size did not differ between FVB and A17.1, but with the 1C protocol the average transcript size was significantly smaller in A17.1 (Fig. 2A). Same as with the 7C (standard) protocol also with the 1C protocol an APA-shift to proximal was prominent (Fig. 2B). Results from permuted half split by shortening of extension of the 3’UTR were similar between 7C and 1C (Fig. S3). Since our APA-shift calculation depends on a split of the 3’UTR in two we expect a correlation with the 3’UTR length, which was confirmed by a linear regression analysis (Fig. S2D). Strikingly, a stronger correlation was found with the (Fig. S2D). Together, the 1C protocol could be more beneficial for APA-shift at the 3’UTR.

**Figure 2.**
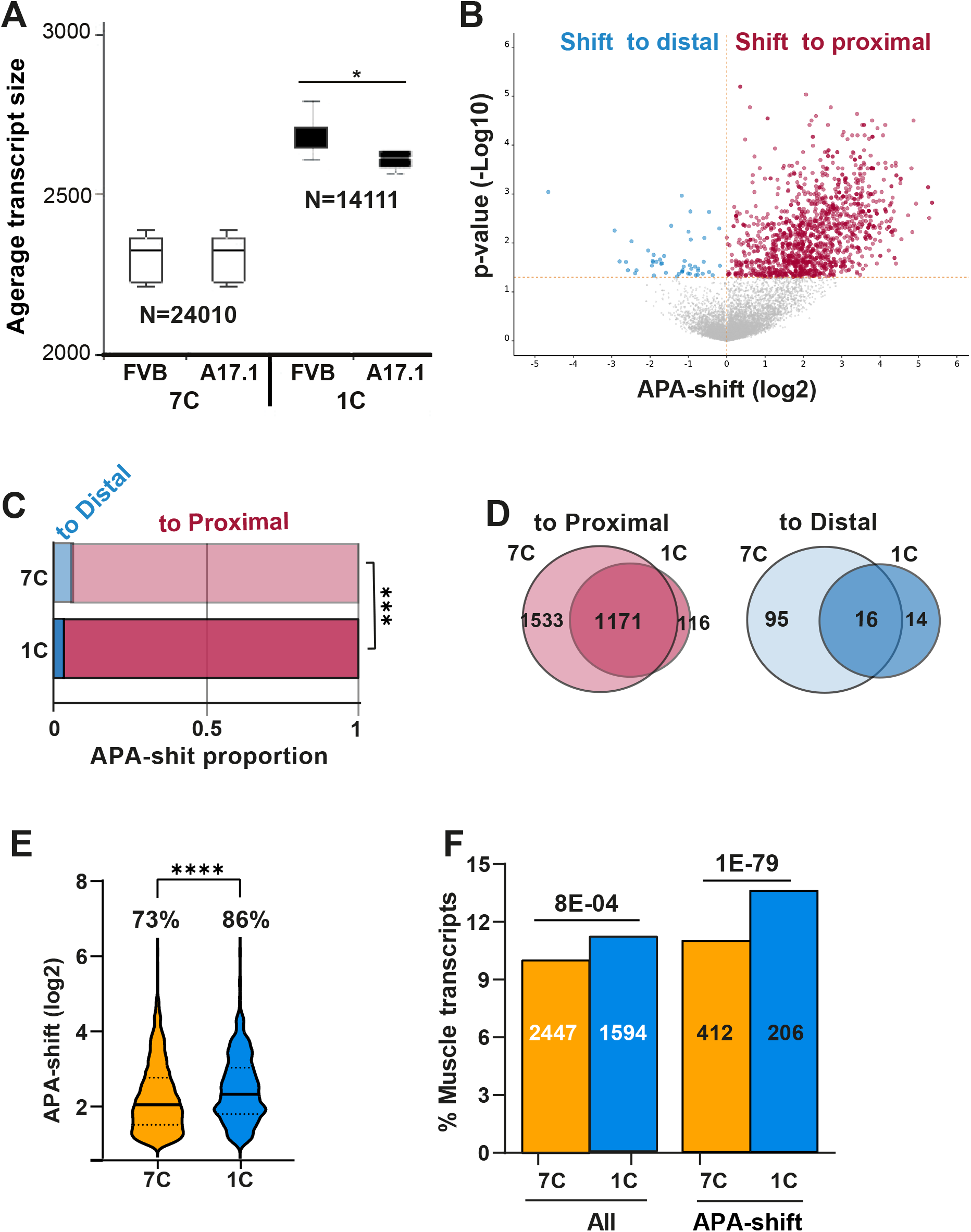
A comparison between 7C and 1C protocols and APA-shift results. **A.** Boxplot shows average transcript size in 7C or 1C libraries in FVB and A17.1 muscles. Number of transcripts (N=) is depicted. Average is from N=5. A significant difference between FVB and A17.1 assessed by the student’s t-test (asterisk indicates p<0.05). **B.** Volcano plot of APA-shift in A17.1 vs. FVB mouse in the 1C RNAseq. Mean -log fold change and *P-*values are from N=5 per genotype. Transcripts with a shift to proximal are highlighted in red and a shift to distal in blue. **C.** Proportions of APA-shift transcripts (p<0.05 FDR) in 7C and 1C RNAseq protocols. A shift to distal is depicted in blue and to proximal in red. Statistical difference was assessed with a chi-square test (*** p<0.0001). **D.** Venn diagrams show the overlap APA-shift (p<0.05 FDR) transcripts between the 7C and the 1C data sets. A shift to proximal in is red and a shift to distal in blue. **E.** Violin plot shows APA-shift (log 2) of the most prominent APA-shift transcripts (p<0.05 FDR and fold change >1) in 7C and 1C data sets. The percentage of most prominent transcripts from APA-shift transcripts is depicted. Statistical significance was assessed with Anova (**** p<0.00001 **F.** Bar chart shows the percentage of muscle genes encoded transcripts in the entire library (all transcripts) and the APA-shift transcripts in 7C and 1C protocols. The numbers of muscle transcripts is depicted in the bar. A statistical difference was assessed with a chi-square test, is depicted above the bars.

To further elucidate the differences between 1C and 7C protocol we compared APA-shift directions. The proportion of transcripts with a shift to distal was significantly smaller in 1C compared with the 7C protocol (Fig. 2C). Moreover, the overlap of transcripts between 1C and 7C was very high for the shift to proximal was 91%, but only 53% for the transcripts with a shift to distal (Fig. 2D). To further investigate differences between 1C and 7C protocols, we focused on the prominent shift to proximal transcripts, defined as APA-shift >1 (log2), as some of the shift to distal transcripts could be false positives. The mean APA-shift value and the percentage of a prominent shift to proximal transcripts were higher in 1C compared to 7C (Fig. 2E).

As the A17.1 show muscle pathology, therefore we investigated if APA-shift of muscle transcripts differed between 7C and 1C. The proportion of muscle transcripts with 3’UTR was higher 1C, moreover, muscle transcripts with APA-shift were more abundant in the 1C protocol (Fig. 2F). This indicates that the 1C protocol is beneficial for muscle transcripts. All subsequent analyses were carried out with the 1C dataset.

### Reduced levels of *Pabpn1* isoforms correlate with APA-shift in A17.1

In A17.1 the expanded PABPN1 cDNA is overexpressed suggesting that expression of the expanded PABPN1 leads to APA-shift. PABPN1 overexpression was confirmed by analysis of sequencing reads in the last exon (Fig 3A). However, sequencing reads at the 3’UTR was slightly lower in the A17.1 (Fig. 3A). The last exon is common between the endogenous Pabpn1 and the expanded PABPN1 transgene but reads from the 3’UTR are only from endogenous *Pabpn1* because the transgene lacks *Pabpn1* 3’UTR. In mice, *Pabpn1* has eight annotated transcript isoforms, but only five isoforms were found in the 1C dataset (Table S5). Levels of isoforms −201, −208, and −209 were significantly lower in A17.1 (Fig. 3B), but expression levels of isoforms −202 and −207 were higher in A17.1 (Fig. 3B and Table S5). The isoforms −201, −202, −207, and −208 encode for a protein containing RNA binding motifs, but the isoform 208 lacks the first exon, containing the polyAla tract (Fig. 3C). The isoform 209 is a non-coding transcript. Considering the differences in expression levels of the five isoforms we assessed a correlation between each isoform and APA-shift values. A negative correlation between isoforms 201, 208, and 209 expression levels and proximal APA-shift indicated that reduced levels in A17.1 correlates with APA-shift to proximal (Fig. 3D). Over 70% of the transcripts with proximal APA-shift showed a significant correlation with isoforms −201, −208 whereas the correlation with a shift to distal was lower (Fig. 3D). In contrast to Isoforms −201 and −208, a weaker positive correlation was found between isoform-207 and an APA-shift to proximal (Fig. 3C). Isoform-202 showed a low correlation with an APA-shift to proximal (Fig. 3D). Interestingly, reduced levels of isoform-209 showed the highest correlation with APA-shift, as over 90% of APA-shift transcripts showed a significant correlation (Fig. 3D). The correlation between reduced levels of PABPN1 isoforms and APA-shift agrees with APA site utilization in PABPN1 knockout cells [19]. Together, our studies here suggests that reduced Pabpn1 level is the cause for APA-shift in A17.1. Yet, with this model we could not resolve the question whether the expanded PABPN1 regulate APA-shifts the same as the normal PABPN1.

**Figure 3.**
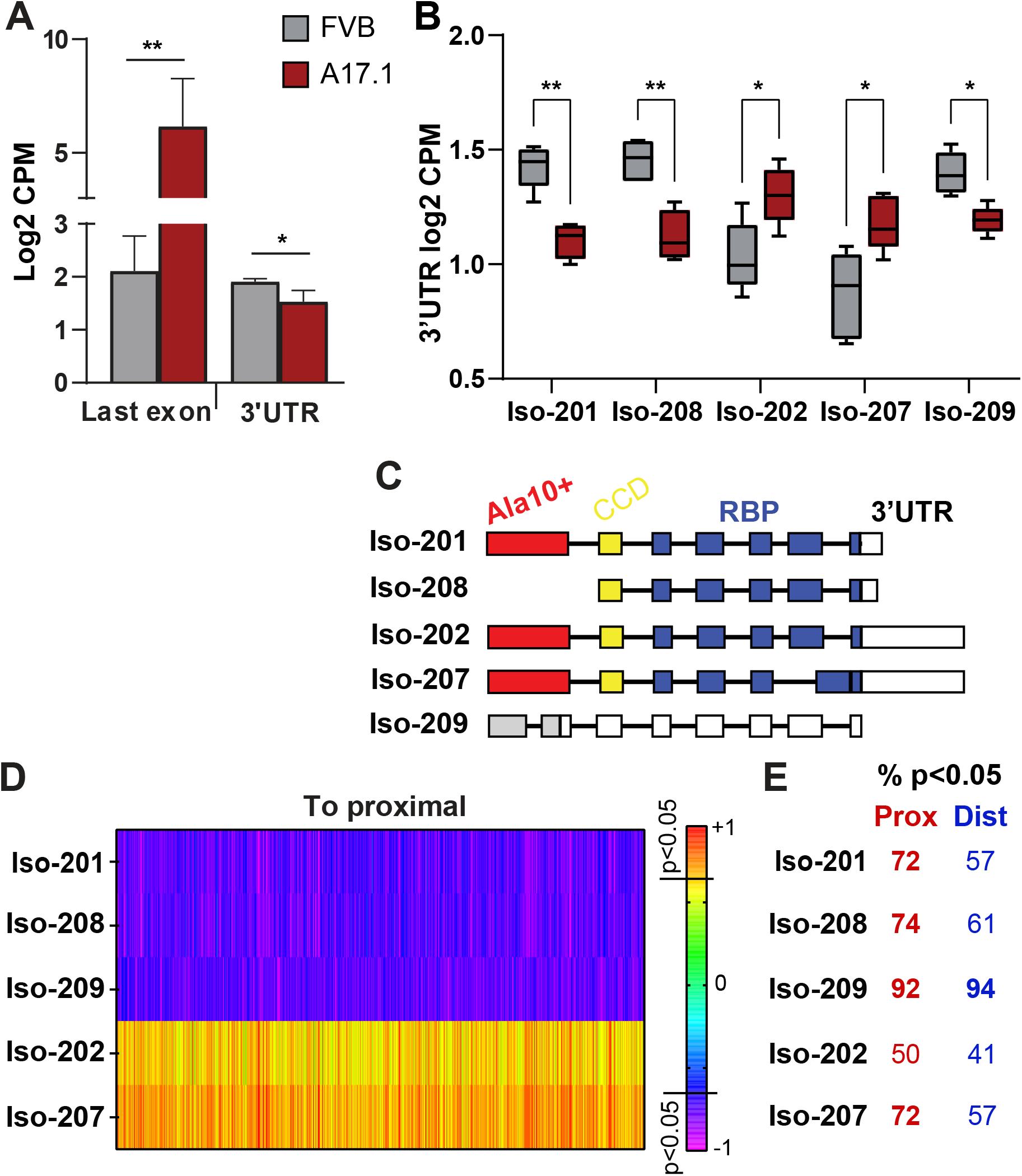
Pabpn1 transcript levels in the A17.1 mouse model are associated with APA-shift. **A.** Bar charts show *Pabpn1* levels measured from 1C RNAseq last axon, representing both endogenous Pabpn1 and the transgene Ala17 PABPN1. Reads from the 3’UTR (sum of reads in all isoforms) represent the endogenous *Pabpn1*. Statistical difference was assessed with a student’s t-test. **B.** Bar chart shows levels of *Pabpn1* transcript isoforms in FVB and A17.1. Statistical difference was assessed with *anova,* adjusted to multiple correction. **C.** A schematic presentation of *Pabpn1* isoforms, the 1st exon encoding for poly Ala is depicted in red, exon encoding for the coil-coil domain is in yellow, and the exons encoding for the RNA binding domain are in blue. The 3’UTR is depicted in white. Isoform 209 is annotated as nonsense mediated decay. **D.** Heatmap of Spearman correlation (r) between APA-shift ratio (log 2) in the APA-shift significant transcripts and Pabpn1 isoforms (-Iso) expression levels (log2). correlation analysis was carried out for transcripts with APA-shift ‘to proximal’ or ‘to distal’ separately, for all five Pabpn1 isoforms. The percentage of transcripts with significant correlation (, p<0.05) is depicted right to the heatmap.

### Low expression levels of expanded PABPN1 does not induce APA-shift

To investigate if expression of the expanded PABPN1 causes APA-shift we utilized a previously reported mouse muscle cell model for OPMD [29]. In this cell model, the expanded PABPN1 (A17) and the normal allele (A10) were over-expressed at similar levels in differentiated muscle cells [29]. PABPN1 over-expression was at low levels, driving only limited aggregation in myonuclei [29]. Therefore, this cell model is a relevant model to assess the effect of the expanded PABPN1 on PABPN1 function, independent of its aggregation. Employing the 1C protocol on RNA from A10 and A17, similar expression levels of PABPN1 transgene alleles were found (Fig. 4A). In contrast to A17.1 mouse model, APA-shift was not genome-wide, and a prominent shift to proximal was not found in A17 cells (Fig. 4B). To validate these results, we repeated the entire procedure using the 7C protocol, resulting in similar low APA-shift as in C1 (Fig. S4). Moreover, with both 1C and 7C protocols the expression of A17 did not lead to a prominent shift to proximal (Fig. S4). This demonstrate that in A17 cell model APA is not activated genome wide.

**Figure 4.**
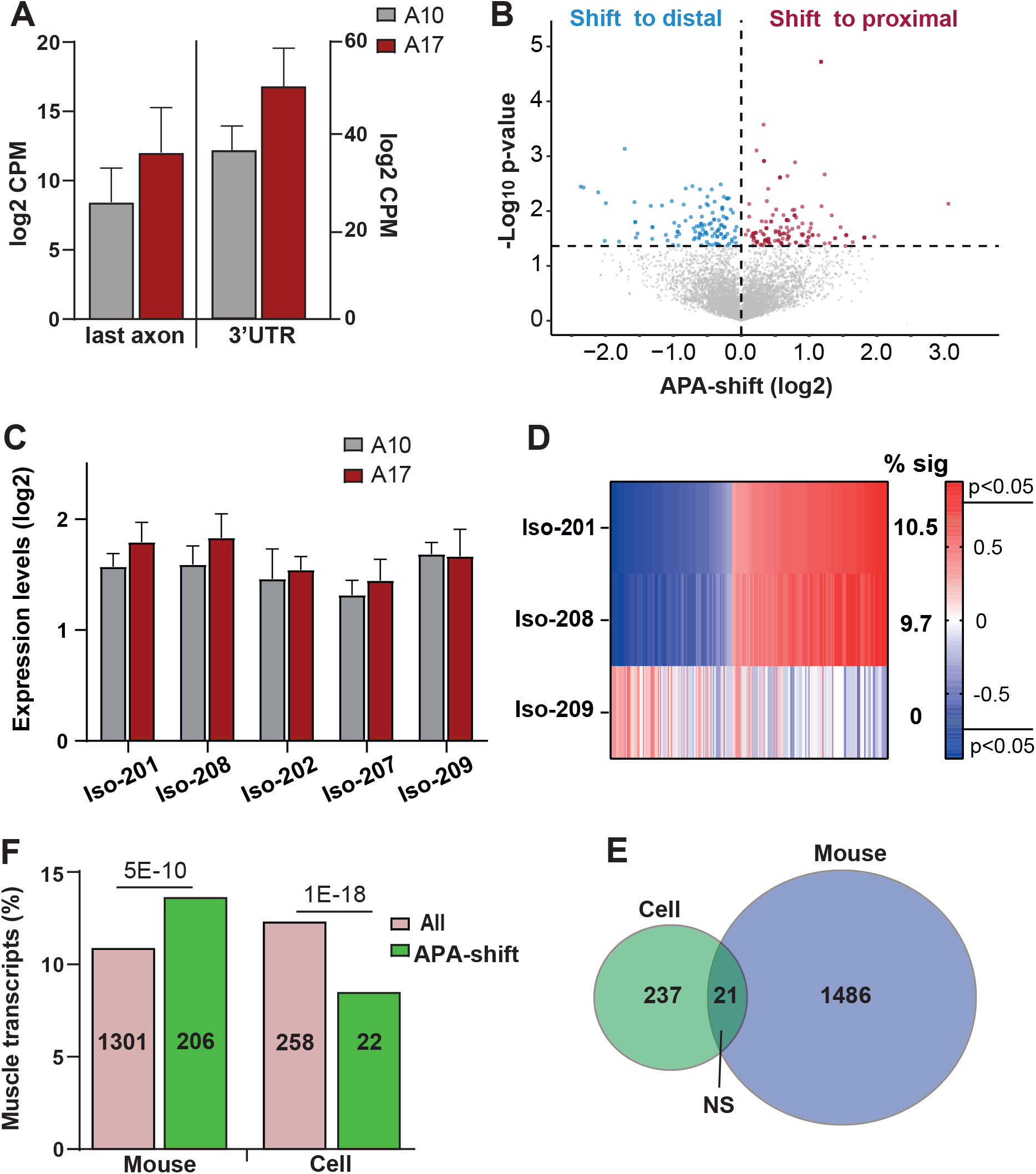
APA-shift in differentiated (fused) A17 muscle cell model is not associated with PABPN1 levels. **A.** Bar charts show *Pabpn1* levels measured from 1C RNAseq last axon, representing both endogenous Pabpn1 and the transgenes Ala17 or Ala10 PABPN1. Reads from the 3’UTR (sum of reads in all isoforms) represent the endogenous *Pabpn1*. Statistical difference was assessed with a student’s t-test. **B.** Volcano plot of APA-shift in A17 vs. A10 muscle cells in the 1C RNAseq. Mean -log fold change and *P-*values are from N=3 per genotype. Transcripts with a switch to proximal are highlighted in red and a switch to distal in blue. **C.** Bar chart shows levels of *Pabpn1* transcripts isoforms expression levels in A10 and A17 muscle cell model. Reads count are from the 3’-UTR. Statistical difference was assessed with *anova,* adjusted to multiple correction. **D.** Heatmap of Spearman correlation (r) between APA-shift ratio (log 2) in the APA-shift significant transcripts and Pabpn1 isoforms (-Iso) expression levels (log2). Correlation analysis was carried out on transcripts with significant APA-shift for three Pabpn1 isoforms. The percentage of transcripts with significant correlation (, p<0.05) is depicted right to the heatmap. **E.** Bar chart shows the percentage of the transcripts encoding for muscle genes in the entire library (all transcripts) and the APA-shift transcripts in the OPMD mouse and cell datasets. Numbers of transcripts is depicted in each bar. Statistical difference was assessed with a chi-square test. **F.** Venn diagram shows transcript overlap for APA-shift transcripts in cell and mouse OPMD models. Statistical significance of overlap was assessed with a chi-square test, NS = not significant.

Interestingly, the expression of the expanded PABPN1 did not cause a reduction in endogenous Pabpn1 expression levels (Fig. 4A). Only isoforms −201 and −208 showed higher levels in A17, but the fold change was small (1.7 folds) (Fig. 4C). A correlation between Isoforms −201 or −208 expression levels and APA-shift was minimal, with only 10% of transcripts with APA-shift showed a significant correlation (Fig. 4D). In contrast to the results in A17.1, isoform-209 did not correlate with APA-shift (Fig. 4D).

Since APA-shift in muscle transcripts was prominent in A17.1 tibialis anterior muscle, we assessed if in the cell model muscle transcripts with APA-shift are also enriched. The percentage of muscle transcripts in the 1C dataset was similar in mice and muscle cell lines (Fig. 4F). However, in the A17 muscle cell culture the proportion of muscle transcripts with APA-shift was significantly lower than the proportion of muscle transcripts in the entire dataset (Fig. 4E). In agreement with little similarities between A17.1 and A17 effect on APA-shift, an overlap between A17 and A17.1 of transcripts with APA-shift was insignificant (Fig. 4F). Together, the results here indicate that expression of the expanded PABPN1 does not cause APA-shift.

### APA-shift in OPMD is enriched in muscle-associated pathways

So far, APA as a readout for PABPN1 function was investigated in OPMD models, but not in OPMD patients. We applied the 1C protocol with the APA-shift analysis on RNA from OPMD (N=10) and control (N=8) vastus lateralis muscle. To our surprise, APA-shift was not massive in OPMD, and a prominent shift to proximal was not found (Fig. 5A). Visualization of reads peak at the 3’UTR, agreed with genomic localization of reads’ peak in using human samples PAS-seq or the poly(A) database (Fig. S4). A PCA of APA-shift values in A17.1 showed a clear separation between control and OPMD model, whereas in humans and the cell model a separation based on genotype was not found (Fig. S4). Despite the low number of transcripts with APA-shift in OPMD enrichment analysis showed an exclusive enrichment for ‘myogenesis’ (Fig. 5B), with most of the muscle transcripts showing a shift to proximal (Table S5). We then assessed an overlap between the muscle genes with APA-shift in OPMD and A17.1. A significant overlap was found at gene level, and most of genes showed an APA-shift to proximal (Fig. 5B, and Table S5). The similarity between OPMD and A17.1 APA-shift was also found in *gene ontology* (GO) enrichment analysis. Similar terms related to muscle pathology were enriched in OPMD and A17.1 (Fig. 5D).

**Figure 5.**
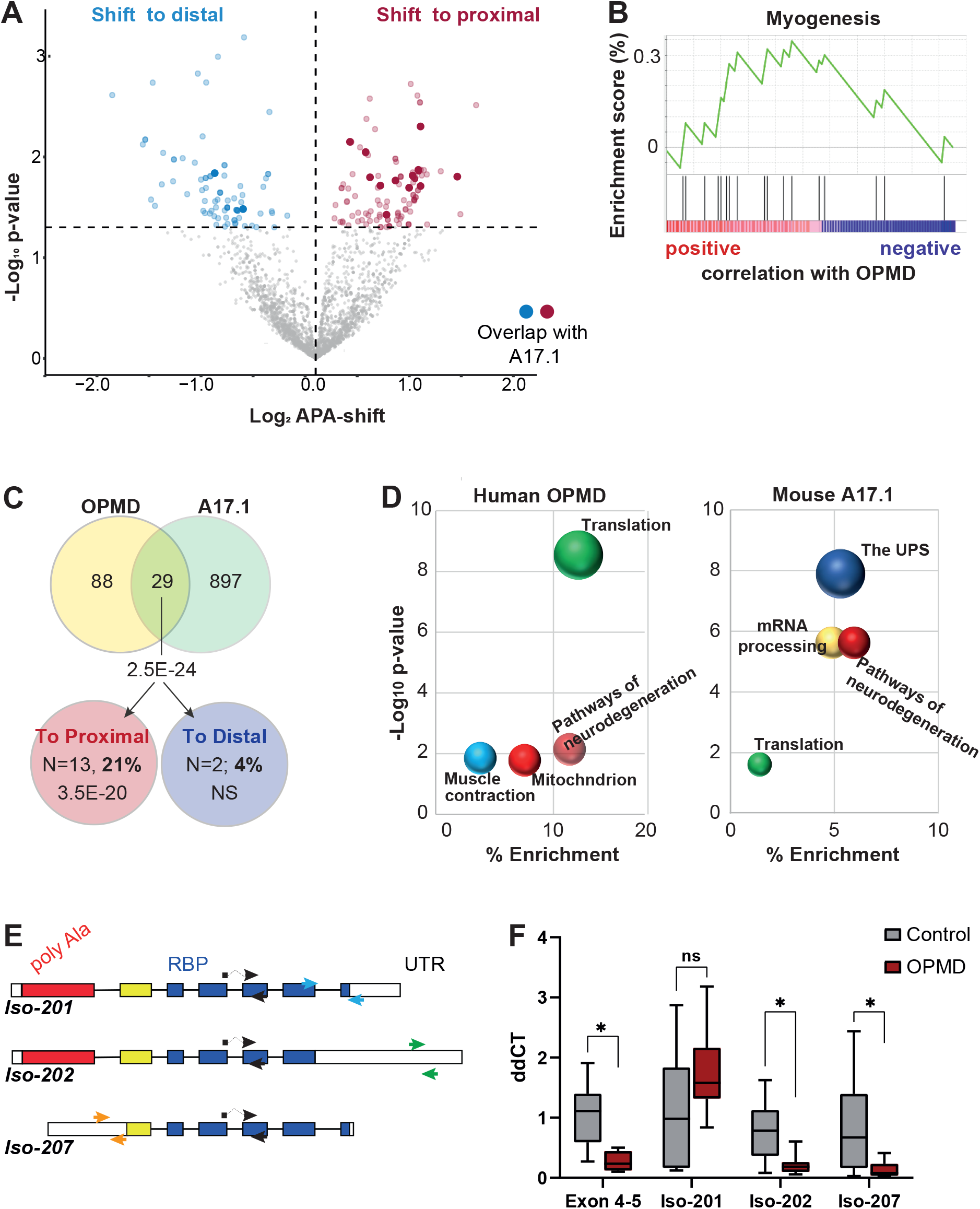
APA shift in OPMD muscles. **A.** Volcano plot of APA-shift in patients with OPMD vs. control’s *vastus lateralis* in 1C RNAseq. Mean -log fold change and *P-*values are from N=10 and 8 per genotype, respectively. Transcripts with a switch to proximal are highlighted in dark red and a switch to distal in dark blue. **B.** Enrichment analysis of OPMD APA-shift transcripts. Visualization of GSEA Enrichment. Only ‘Myogenesis’ was found significant, and most transcripts showed positive correlation with OPMD. **C.** Venn diagram shows the overlap of genes with significant APA-shift between OPMD and A17.1 in yellow and green, respectively, and among genes with proximal (red) or distal (blue) APA-shift. The overlap significance was assessed with a chi-square test. OPMD APA-shift genes overlapping with A17.1 and enriched in skeletal muscles are depicted. **D.** Dot plot visualization of functional term enrichment for the proximal APA-shift genes in OPMD and in A17.1. Redundancy was removed using annotation clustering, the most significant term within an enrichment cluster is depicted. Similar terms are marked with the same color. **E.** A schematic presentation of *PABPN1* isoforms −201, −202 and −207. Exons are depicted in full boxes and introns in dashed lines. The first exon encoding for polyAla is depicted in red, exon encoding for the coil-coil domain is in yellow, and the exons encoding for the RNA binding domain are in blue. UTR regions are depicted in white. Primer sets that were used for RT-qPCR are denoted with arrows: exon 4-5 in black arrows; Iso-001 in blue; Iso-002 in green and Iso-004 in orange. **F.** Bar chart shows normalized ddCT values of *PABPN1* isoforms in control and OPMD. Statistical difference was assessed with *anova*: ** p<0.0005; * p<0.05.

In a previous study we showed reduced *PABPN1* levels in OPMD[2], here we investigated whether *PABPN1* isoforms might impact APA-shift in OPMD. In humans, the three isoforms −001 (ENST00000216727), −003 (ENST00000397276) and −004 (ENST00000556821) are the most abundant in skeletal muscles (https://www.proteinatlas.org/ and [41]). The isoforms −001 and −002 encode for the full length PABPN1 but differ in UTR sequences (Fig. 5A). Isoform-004 lacks the majority of the first exon, including the polyAla expansion (Fig. 5A). Consistent with our previous study, also in this study we detected reduced *PABPN1* levels in OPMD but using primers specific to iso-001 or iso-004 reduced PABPN1 levels was specific to Iso-004 (Fig. 5B). This suggests that APA-shift in OPMD is driven by reduced iso-004 levels.

## Discussion

Genome-wide alterations in transcripts expression levels due to APA have been linked to a range of pathological conditions [15]. Albeit cell types can be distinguished by patterns of APA marking cell specificity [34], in pathological conditions, APA has been found in a wide range of tissues including different cancer types [8, 14, 23, 45], heart [9], brain [10], cells of the immune response [20]. Here we show APA-shift in skeletal muscles from OPMD patients. Thus, our study corroborates with other studies suggesting that APA as an important regulatory layer in human pathologies.

Identification of APA site utilization requires specific RNA sequencing protocols, such as PAS-seq, which were instrumental to distinguish between the proximal and distal PASs [12, 44]. In OPMD mouse model that proximal PAS mostly represent APA sites [12, 19]. However, PAS-seq is costly and hence less suitable for patient material. Computational pipelines were developed to accurately identify alteration in APA and PAS site utilization [16, 21, 43]. However, since PAS utilization shows cell type and species specify [34], application of this procedure in less studied cells, such as muscle cells could be tricky [24]. We developed the C1 protocol and in combination with the APA-shift computational pipeline it reports PABPN1 function in a single quantitative readout. Compared to the standard protocol, in the 1C protocol, RNA dynamics are closer to the in vivo situation. We show that the 1C protocol is more suitable for muscle transcripts and detecting APA-shifts. Yet, the 1C has a few cons: its sequencing depth is not as deep as the standard protocol and is more dependent on a similar amount and quality of RNA input across samples. The latter could be a challenge for tissues that are stored for a long time. The APA-shift pipeline is independent of PAS genomic information, and hence is only an estimation for PAS utilization. Yet, APA-shift and PAS-seq in the mouse OPMD model showed nearly 80% transcript overlap indicating APA-shift reliability. Our pipeline here is simple, contains fewer assumptions, and can be applied to existing RNA sequencing data to assess APA-shift.

Although APA site utilization is regulated by multiple nuclear RNA binding proteins, reduced PABPN1 levels have been implicated in APA site utilization in different cancers [8, 18]. Reduced PABPN1 levels leads to muscle wasting[27, 32], and have been also found in OPMD patients [2, 33]. Although genome-wide APA has been previously demonstrated in a mouse model for OPMD that overexpress the expanded PABPN1 [12, 19], in OPMD muscles we found APA-shift mainly in muscle transcripts. The limited APA-shift we found in OPMD could be attributed to the C1 protocol that was not genome-wide in RNA from patients’ muscles. Also, in lung cancer cells APA was found in a small number of genes regulating cell cycle [18]. Together, it is possible that APA is initially found in a small subset of transcripts that may drive cell damage. In skeletal muscles the muscle transcripts with APA-shift could contribute to muscle weakness and atrophy.

Our results here show that the expression of the pathogenic, alanine-expanded, PABPN1 does not cause APA-shift. Instead, reduced expression levels of a subset of PABPN1 isoforms correlate with APA-shift in both A17.1 mouse model and in OPMD. It is possible that (over) expression of the expanded PABPN1 leads to reduced expression of the normal transcript isoforms. It was previously suggested by others and us that PABPN1 aggregation leads reduced levels of the functional PABPN1 [4, 30], but the exact molecular mechanism is unknown. In mouse the non-coding *Pabpn1* isoform-209, was expressed at levels close to the protein-coding isoforms and showed the strongest correlation with APA-shift. Non-coding RNAs can regulate RNA expression levels [35]. A model for PABPN1 self-regulation mechanism has been proposed, where high PABPN1 protein expression levels lead to reduced expression via mRNA decay [5]. The human non-coding PABPN1 isoforms are not abundant in skeletal muscles (https://www.proteinatlas.org). OPMD is mostly considered as a gain of function disease due to the presence of nuclear aggregates [22]. Our study suggests that PABPN1 loss of function also contributes to OPMD. Interestingly, we identify that levels of the isoform lacking the alanine expansion region are reduced on OPMD. The role of the truncated isoform is unknown, but potentially it could open specific therapeutical options for OPMD.

To summarize: the combination of 1C protocol and APA-shift quantification allowed us, for the first time, to report APA-shift in OPMD patients. APA-shift in OPMD is predominantly enriched in muscle genes and genes affecting neurodegeneration pathways. The mutated PABPN1 does not affect APA-shift, suggesting its expression does not compromise PABPN1 function. Instead, lower expression of PABPN1 isoforms could compromise PABPN1 function.

## Data and code availability

APA-shift results in all models and in OPMD are provided in <u>Table S3</u>. Individual sequencing data are not available due to participants privacy.

Codes that were used in this study are available upon request.

## Data Availability

All data produced in the present study are available upon reasonable request to the authors

## Acknowledgement

This study is supported by the Human Genetics NGS funding, and by the kind support of argenx. We thank Peter A.C, ‘t Hoen (Department of Medical BioSciences, Radboud university medical center, Nijmegen, The Netherlands) for critical comments and useful suggestions on the manuscript.

## References

1 Andreassi C, Riccio A (2009) To localize or not to localize: mRNA fate is in 3ʹUTR ends. Trends in Cell Biology 19: 465–474 Doi https://doi.org/10.1016/j.tcb.2009.06.001

2 Anvar SY, Raz Y, Verway N, van der Sluijs B, Venema A, Goeman JJ, Vissing J, van der Maarel SM, t Hoen PA, van Engelen BG et al (2013) A decline in PABPN1 induces progressive muscle weakness in oculopharyngeal muscle dystrophy and in muscle aging. Aging (Albany NY) 5: 412–426 Doi 10.18632/aging.100567

3 Bagnoli JW, Ziegenhain C, Janjic A, Wange LE, Vieth B, Parekh S, Geuder J, Hellmann I, Enard W (2018) Sensitive and powerful single-cell RNA sequencing using mcSCRB-seq. Nature Communications 9: 2937 Doi 10.1038/s41467-018-05347-6

4 Banerjee A, Apponi LH, Pavlath GK, Corbett AH (2013) PABPN1: molecular function and muscle disease. Febs j 280: 4230–4250 Doi 10.1111/febs.12294

5 Bergeron D, Pal G, Beaulieu YB, Chabot B, Bachand F (2015) Regulated Intron Retention and Nuclear Pre-mRNA Decay Contribute to PABPN1 Autoregulation. Mol Cell Biol 35: 2503–2517 Doi 10.1128/MCB.00070-15

6 Brais B, Bouchard JP, Xie YG, Rochefort DL, Chrétien N, Tomé FM, Lafrenière RG, Rommens JM, Uyama E, Nohira Oet al (1998) Short GCG expansions in the PABP2 gene cause oculopharyngeal muscular dystrophy. Nat Genet 18: 164–167 Doi 10.1038/ng0298-164

7 Chang JW, Yeh HS, Yong J (2017) Alternative Polyadenylation in Human Diseases. Endocrinol Metab (Seoul) 32: 413–421 Doi 10.3803/EnM.2017.32.4.413

8 Chen L, Dong W, Zhou M, Yang C, Xiong M, Kazobinka G, Chen Z, Xing Y, Hou T (2023) PABPN1 regulates mRNA alternative polyadenylation to inhibit bladder cancer progression. Cell & Bioscience 13: 45 Doi 10.1186/s13578-023-00997-6

9 Creemers EE, Bawazeer A, Ugalde AP, van Deutekom HW, van der Made I, de Groot NE, Adriaens ME, Cook SA, Bezzina CR, Hubner N et al (2016) Genome-Wide Polyadenylation Maps Reveal Dynamic mRNA 3’-End Formation in the Failing Human Heart. Circ Res 118: 433–438 Doi 10.1161/circresaha.115.307082

10 Cui Y, Arnold FJ, Peng F, Wang D, Li JS, Michels S, Wagner EJ, La Spada AR, Li W (2023) Alternative polyadenylation transcriptome-wide association study identifies APA-linked susceptibility genes in brain disorders. Nat Commun 14: 583 Doi 10.1038/s41467-023-36311-8

11 Datlinger P, Rendeiro AF, Boenke T, Senekowitsch M, Krausgruber T, Barreca D, Bock C (2021) Ultra-high-throughput single-cell RNA sequencing and perturbation screening with combinatorial fluidic indexing. Nature Methods 18: 635–642 Doi 10.1038/s41592-021-01153-z

12 de Klerk E, Venema A, Anvar SY, Goeman JJ, Hu O, Trollet C, Dickson G, den Dunnen JT, van der Maarel SM, Raz V et al (2012) Poly(A) binding protein nuclear 1 levels affect alternative polyadenylation. Nucleic Acids Res 40: 9089–9101 Doi 10.1093/nar/gks655

13 de Leeuw RH, Garnier D, Kroon R, Horlings CGC, de Meijer E, Buermans H, van Engelen BGM, de Knijff P, Raz V (2019) Diagnostics of short tandem repeat expansion variants using massively parallel sequencing and componential tools. Eur J Hum Genet 27: 400–407 Doi 10.1038/s41431-018-0302-4

14 Goering R, Engel KL, Gillen AE, Fong N, Bentley DL, Taliaferro JM (2021) LABRAT reveals association of alternative polyadenylation with transcript localization, RNA binding protein expression, transcription speed, and cancer survival. BMC Genomics 22: 476 Doi 10.1186/s12864-021-07781-1

15 Gruber AJ, Zavolan M (2019) Alternative cleavage and polyadenylation in health and disease. Nature Reviews Genetics 20: 599–614 Doi 10.1038/s41576-019-0145-z

16 Ha KCH, Blencowe BJ, Morris Q (2018) QAPA: a new method for the systematic analysis of alternative polyadenylation from RNA-seq data. Genome Biol 19: 45 Doi 10.1186/s13059-018-1414-4

17 Huang DW, Sherman BT, Tan Q, Kir J, Liu D, Bryant D, Guo Y, Stephens R, Baseler MW, Lane HC et al (2007) DAVID Bioinformatics Resources: expanded annotation database and novel algorithms to better extract biology from large gene lists. Nucleic Acids Res 35: W169–175 Doi 10.1093/nar/gkm415

18 Ichinose J, Watanabe K, Sano A, Nagase T, Nakajima J, Fukayama M, Yatomi Y, Ohishi N, Takai D (2014) Alternative polyadenylation is associated with lower expression of PABPN1 and poor prognosis in non-small cell lung cancer. Cancer Sci 105: 1135–1141 Doi 10.1111/cas.12472

19 Jenal M, Elkon R, Loayza-Puch F, van Haaften G, Kühn U, Menzies FM, Oude Vrielink JA, Bos AJ, Drost J, Rooijers K et al (2012) The poly(A)-binding protein nuclear 1 suppresses alternative cleavage and polyadenylation sites. Cell 149: 538–553 Doi 10.1016/j.cell.2012.03.022

20 Jia X, Yuan S, Wang Y, Fu Y, Ge Y, Ge Y, Lan X, Feng Y, Qiu F, Li Pet al (2017) The role of alternative polyadenylation in the antiviral innate immune response. Nat Commun 8: 14605 Doi 10.1038/ncomms14605

21 Long Y, Zhang B, Tian S, Chan JJ, Zhou J, Li Z, Li Y, An Z, Liao X, Wang Yet al (2023) Accurate transcriptome-wide identification and quantification of alternative polyadenylation from RNA-seq data with APAIQ. Genome Res 33: 644–657 Doi 10.1101/gr.277177.122

22 Malerba A, Klein P, Bachtarzi H, Jarmin SA, Cordova G, Ferry A, Strings V, Espinoza MP, Mamchaoui K, Blumen SC et al (2017) PABPN1 gene therapy for oculopharyngeal muscular dystrophy. Nature Communications 8: 14848 Doi 10.1038/ncomms14848

23 Mayr C, Bartel DP (2009) Widespread shortening of 3’UTRs by alternative cleavage and polyadenylation activates oncogenes in cancer cells. Cell 138: 673–684 Doi 10.1016/j.cell.2009.06.016

24 Mei H, Boom J, El Abdellaoui S, Abdelmohsen K, Munk R, Martindale JL, Kloet S, Kielbasa SM, Sharp TH, Gorospe Met al (2022) Alternative Polyadenylation Utilization Results in Ribosome Assembly and mRNA Translation Deficiencies in a Model for Muscle Aging. J Gerontol A Biol Sci Med Sci 77: 1130–1140 Doi 10.1093/gerona/glac058

25 Mitschka S, Mayr C (2022) Context-specific regulation and function of mRNA alternative polyadenylation. Nature Reviews Molecular Cell Biology 23: 779–796 Doi 10.1038/s41580-022-00507-5

26 Nishtala S, Neelamraju Y, Janga SC (2016) Dissecting the expression relationships between RNA-binding proteins and their cognate targets in eukaryotic post-transcriptional regulatory networks. Scientific Reports 6: 25711 Doi 10.1038/srep25711

27 Olie CS, Riaz M, Konietzny R, Charles PD, Pinto-Fernandez A, Kiełbasa SM, Aartsma-Rus A, Goeman JJ, Kessler BM, Raz V (2019) Deacetylation Inhibition Reverses PABPN1-Dependent Muscle Wasting. iScience 12: 318–332 Doi 10.1016/j.isci.2019.01.024

28 Raz V, Buijze H, Raz Y, Verwey N, Anvar SY, Aartsma-Rus A, van der Maarel SM (2014) A novel feed-forward loop between ARIH2 E3-ligase and PABPN1 regulates aging-associated muscle degeneration. Am J Pathol 184: 1119–1131 Doi 10.1016/j.ajpath.2013.12.011

29 Raz V, Routledge S, Venema A, Buijze H, van der Wal E, Anvar S, Straasheijm KR, Klooster R, Antoniou M, van der Maarel SM (2011) Modeling oculopharyngeal muscular dystrophy in myotube cultures reveals reduced accumulation of soluble mutant PABPN1 protein. Am J Pathol 179: 1988–2000 Doi 10.1016/j.ajpath.2011.06.044

30 Raz Y, Raz V (2014) Oculopharyngeal muscular dystrophy as a paradigm for muscle aging. Front Aging Neurosci 6: 317 Doi 10.3389/fnagi.2014.00317

31 Ren F, Zhang N, Zhang L, Miller E, Pu JJ (2020) Alternative Polyadenylation: a new frontier in post transcriptional regulation. Biomark Res 8: 67 Doi 10.1186/s40364-020-00249-6

32 Riaz M, Raz Y, van Putten M, Paniagua-Soriano G, Krom YD, Florea BI, Raz V (2016) PABPN1-Dependent mRNA Processing Induces Muscle Wasting. PLoS Genet 12: e1006031 Doi 10.1371/journal.pgen.1006031

33 Roth F, Dhiab J, Boulinguiez A, Mouigni H-R, Lassche S, Negroni E, Muraine L, Marhic A, Oliver A, Lainé Jet al (2022) Assessment of PABPN1 nuclear inclusions on a large cohort of patients and in a human xenograft model of oculopharyngeal muscular dystrophy. Acta Neuropathologica 144: 1157–1170 Doi 10.1007/s00401-022-02503-7

34 Shulman ED, Elkon R (2019) Cell-type-specific analysis of alternative polyadenylation using single-cell transcriptomics data. Nucleic Acids Res 47: 10027–10039 Doi 10.1093/nar/gkz781

35 Statello L, Guo C-J, Chen L-L, Huarte M (2021) Gene regulation by long non-coding RNAs and its biological functions. Nature Reviews Molecular Cell Biology 22: 96–118 Doi 10.1038/s41580-020-00315-9

36 Subramanian A, Tamayo P, Mootha VK, Mukherjee S, Ebert BL, Gillette MA, Paulovich A, Pomeroy SL, Golub TR, Lander ES et al (2005) Gene set enrichment analysis: a knowledge-based approach for interpreting genome-wide expression profiles. Proc Natl Acad Sci U S A 102: 15545–15550 Doi 10.1073/pnas.0506580102

37 Tanguay RL, Gallie DR (1996) Translational efficiency is regulated by the length of the 3’ untranslated region. Mol Cell Biol 16: 146–156 Doi 10.1128/mcb.16.1.146

38 Tavanez JP, Calado P, Braga J, Lafarga M, Carmo-Fonseca M (2005) In vivo aggregation properties of the nuclear poly(A)-binding protein PABPN1. Rna 11: 752–762 Doi 10.1261/rna.7217105

39 Tian B, Manley JL (2017) Alternative polyadenylation of mRNA precursors. Nature Reviews Molecular Cell Biology 18: 18–30 Doi 10.1038/nrm.2016.116

40 Trollet C, Anvar SY, Venema A, Hargreaves IP, Foster K, Vignaud A, Ferry A, Negroni E, Hourde C, Baraibar MA et al (2010) Molecular and phenotypic characterization of a mouse model of oculopharyngeal muscular dystrophy reveals severe muscular atrophy restricted to fast glycolytic fibres. Hum Mol Genet 19: 2191–2207 Doi 10.1093/hmg/ddq098

41 Uhlén M, Fagerberg L, Hallström BM, Lindskog C, Oksvold P, Mardinoglu A, Sivertsson Å, Kampf C, Sjöstedt E, Asplund Aet al (2015) Proteomics. Tissue-based map of the human proteome. Science 347: 1260419 Doi 10.1126/science.1260419

42 Wilkins OG, Capitanchik C, Luscombe NM, Ule J (2021) Ultraplex: A rapid, flexible, all-in-one fastq demultiplexer. Wellcome Open Res 6: 141 Doi 10.12688/wellcomeopenres.16791.1

43 Ye C, Long Y, Ji G, Li QQ, Wu X (2018) APAtrap: identification and quantification of alternative polyadenylation sites from RNA-seq data. Bioinformatics 34: 1841–1849 Doi 10.1093/bioinformatics/bty029

44 Yoon Y, Soles LV, Shi Y (2021) PAS-seq 2: A fast and sensitive method for global profiling of polyadenylated RNAs. Methods Enzymol 655: 25–35 Doi 10.1016/bs.mie.2021.03.013

45 Yuan F, Hankey W, Wagner EJ, Li W, Wang Q (2021) Alternative polyadenylation of mRNA and its role in cancer. Genes & Diseases 8: 61–72 Doi https://doi.org/10.1016/j.gendis.2019.10.011

