## Supplementary material for "*PABPN1* loss-of-function in oculopharyngeal muscular dystrophy primarily impacts APA-shift in muscle transcripts": Suppl data: Supplementary data.pdf

#### A. Supplementary Tables

**Table S1**: Gender and age information of OPMD patients and controls whose RNA from vastus lateralis muscle was used in this study.

| OPMD | | | Control | | |
|---|---|---|---|---|---|
| Sample nr | **Age range** | **Gender** | Sample nr | **Age range** | **Gender** |
| 1 | 60-69 | f | 3 | 20-29 | m |
| 2 | 40-49 | m | 4 | 60-69 | m |
| 3 | 50-59 | m | 5 | 60-69 | f |
| 4 | 80-89 | f | 6 | 80-89 | f |
| 5 | 40-49 | f | 7 | 30-39 | m |
| 6 | 50-59 | f | 8 | 40-49 | f |
| 7 | 40-49 | f | 11 | 80-49 | f |
| 8 | 70-79 | m | 12 | 50-59 | m |
| 9 | 60-69 | f | | | |
| 10 | 40-49 | f | | | |

**Table S2: Data sets description**

| **A.** **Experiment** | **Sample nr.** | **Library prep** | **Transcript exclusion criteria** | **Transcript nr. (3'UTR)** | **Included transcript nr.** | **Transcript with APA shift** |
|---|---|---|---|---|---|---|
| **Mouse (FVB and A17.1)** | 5 + 5 | 7C | ∑ reads proximal =<10 and | 52218 | 24106 | 3728 |
| | 5 + 5 | 1C | | | 14111 | 1507 |
| **Mouse cell line (A10 and A17)** | 3 + 3 | 7C | | | 23869 | 753 |
| | 3 + 3 | 1C | | | 8278 | 258 |
| **Human (control and OPMD)** | 8 + 10 | 1C | Reads at proximal region in = > 30% of samples | 85619 | 3113 | 215 |

| **B.** | **Category** | **Total input reads** | **Unmapped reads** | | **Mapped reads** | | **Reads after UMI based dedup (transcriptome based approach)** | |
|---|---|---|---|---|---|---|---|---|
| **7C** | FVB_Q2 | 12404400 | 337395 | 2.72% | 12067005 | 97.28% | 6939372 | 57.51% |
| | FVB_Q3 | 13815810 | 342575 | 2.48% | 13473235 | 97.52% | 7080216 | 52.55% |
| | FVB_Q4 | 12525017 | 301601 | 2.41% | 12223416 | 97.59% | 8004434 | 65.48% |
| | FVB_Q5 | 13073935 | 320279 | 2.45% | 12753656 | 97.55% | 7985539 | 62.61% |
| | FVB_Q6 | 11424354 | 297712 | 2.61% | 11126642 | 97.39% | 7445899 | 66.92% |
| | A17.1_Q2 | 11229458 | 315988 | 2.81% | 10913470 | 97.19% | 6485672 | 59.43% |
| | A17.1_Q3 | 10951353 | 310997 | 2.84% | 10640356 | 97.16% | 5586852 | 52.51% |
| | A17.1_Q4 | 11602273 | 334719 | 2.88% | 11267554 | 97.12% | 6286475 | 55.79% |
| | A17.1_Q5 | 10327784 | 294344 | 2.85% | 10033440 | 97.15% | 5854875 | 58.35% |
| | A17.1_Q6 | 10357987 | 319769 | 3.09% | 10038218 | 96.91% | 6209008 | 61.85% |

| | | | | | | | | |
|---|---|---|---|---|---|---|---|---|
| | A10_1 | 14786997 | 514065 | 3.48% | 14272932 | 96.52% | 10443091 | 73.17% |
| | A10_2 | 9284162 | 321864 | 3.47% | 8962298 | 96.53% | 6911140 | 77.11% |
| | A10_3 | 9044039 | 292607 | 3.24% | 8751432 | 96.76% | 6568761 | 75.06% |
| | A17_1 | 10643513 | 388054 | 3.65% | 10255459 | 96.35% | 7073487 | 68.97% |
| | A17_2 | 11148411 | 370948 | 3.33% | 10777463 | 96.67% | 8018058 | 74.40% |
| | A17_3 | 11290889 | 341323 | 3.02% | 10949566 | 96.98% | 7653099 | 69.89% |
| **1C** | FVB_Q2 | 1550669 | 475119 | 30.64% | 1075550 | 69.36% | 871531 | 81.03% |
| | FVB_Q3 | 1816421 | 546595 | 30.09% | 1269826 | 69.91% | 960845 | 75.67% |
| | FVB_Q4 | 1635290 | 494250 | 30.22% | 1141040 | 69.78% | 982341 | 86.09% |
| | FVB_Q5 | 2274844 | 696713 | 30.63% | 1578131 | 69.37% | 1316496 | 83.42% |
| | FVB_Q6 | 2268734 | 698919 | 30.81% | 1569815 | 69.19% | 1344212 | 85.63% |
| | A17.1_Q2 | 1418780 | 443039 | 31.23% | 975741 | 68.77% | 791623 | 81.13% |
| | A17.1_Q3 | 932978 | 289091 | 30.99% | 643887 | 69.01% | 498184 | 77.37% |
| | A17.1_Q4 | 1606702 | 483546 | 30.10% | 1123156 | 69.90% | 878087 | 78.18% |
| | A17.1_Q5 | 1716088 | 525516 | 30.62% | 1190572 | 69.38% | 941188 | 79.05% |
| | A17.1_Q6 | 1638100 | 507705 | 30.99% | 1130395 | 69.01% | 930914 | 82.35% |
| | A10_1 | 556663 | 146303 | 26.28% | 410360 | 73.72% | 364145 | 88.74% |
| | A10_2 | 484524 | 131897 | 27.22% | 352627 | 72.78% | 315090 | 89.36% |
| | A10_3 | 355590 | 104663 | 29.43% | 250927 | 70.57% | 223657 | 89.13% |
| | A17_1 | 153917 | 44902 | 29.17% | 109015 | 70.83% | 97100 | 89.07% |
| | A17_2 | 280383 | 78810 | 28.11% | 201573 | 71.89% | 179363 | 88.98% |
| | A17_3 | 530802 | 147575 | 27.80% | 383227 | 72.20% | 338299 | 88.28% |
| **Control** | c11 | 2117733 | 1824795 | 86.17% | 292938 | 13.83% | 92274 | 31.50% |
| | c12 | 4182735 | 3391372 | 81.08% | 791363 | 18.92% | 236127 | 29.84% |
| | c3 | 1840311 | 1558919 | 84.71% | 281392 | 15.29% | 81990 | 29.14% |
| | c4 | 2267725 | 1903233 | 83.93% | 364492 | 16.07% | 84056 | 23.06% |
| | c5 | 1686682 | 1388817 | 82.34% | 297865 | 17.66% | 84010 | 28.20% |
| | c6 | 2198539 | 1861224 | 84.66% | 337315 | 15.34% | 126287 | 37.44% |
| | c7 | 1467447 | 1329313 | 90.59% | 138134 | 9.41% | 70051 | 50.71% |
| | c8 | 3574491 | 3171647 | 88.73% | 402844 | 11.27% | 179056 | 44.45% |
| **OPMD** | p1 | 4236462 | 3618811 | 85.42% | 617651 | 14.58% | 179128 | 29.00% |
| | p2 | 3269827 | 2851317 | 87.20% | 418510 | 12.80% | 140022 | 33.46% |
| | p3 | 2399867 | 2073306 | 86.39% | 326561 | 13.61% | 101192 | 30.99% |
| | p4 | 2868232 | 2414007 | 84.16% | 454225 | 15.84% | 124061 | 27.31% |
| | p5 | 3371018 | 2689161 | 79.77% | 681857 | 20.23% | 176315 | 25.86% |
| | p6 | 1674483 | 1437591 | 85.85% | 236892 | 14.15% | 89196 | 37.65% |
| | p7 | 3700454 | 3243308 | 87.65% | 457146 | 12.35% | 158209 | 34.61% |
| | p8 | 6500730 | 5446068 | 83.78% | 1054662 | 16.22% | 397067 | 37.65% |
| | p9 | 2386840 | 1875333 | 78.57% | 511507 | 21.43% | 159734 | 31.23% |
| | p10 | 2849503 | 2313485 | 81.19% | 536018 | 18.81% | 137917 | 25.73% |

## C.

| | |
|---|---|
| SCIFI_LIG384 | GAGTTCAGACGTGTGCTCTTCCGATCT-NNNNNNNN-AAGTGATTAGCAA-TTTTTTTTTTTTTTTTTTTTTTTTTTTTTTTVN |
| Well barcode | CAAGCAGAAGACGGCATACGAGAT[i7_barcode]GTGACTGGAGTTCAGACGTGTGCTCTTCCGATCT) |

| D. | Forward 5' – 3' | Reverse 5' – 3' |
|---|---|---|
| Exon 4-5<br>ENSG00000100836 | ATGTTGGCAATGTGGACTATG | ACACGGTTGACTGAACCACA |
| Iso-201<br>ENST00000216727 | GTTTTAACAGCAGGCCCCG | TCTTTTTTTCCTCTCTCTCCTCCTAATAC |
| Iso-202<br>ENST00000397276 | CAACAGCCTTGTGGGAGGAT | CAAAAACCTGGGCACCACAC |
| Iso-207<br>ENST00000556821 | CCCGACTGGCTTGATTCGG | CATGCTCGGCCATTTCCCT |
| HPRT1 | TGGTCAGGCAGTATAATCCAAAGA | TCAAATCCAACAAAGTCTGGCTTA |

**A.** Summary of data sets that were used in this study, transcripts' number that was used for APA-shift calculation and the number of significant APA-shift transcripts in each experiment.

**B.** RNAseq quality control in 7C and 1C protocols in OPMD mouse and cell line models, and in human muscles (controls and OPMD).

**C.** Primers used for library preparation.

**D.** Primers used for RT-qPCR

**Table S3: Transcripts with APA-shift in mouse, cell line and human samples.**

**Table S4: List of muscle transcripts that were included in this study.**

**Table S5: expression levels of PABPN1 transcript isoforms in mouse and cell models.**

**Mouse Tibialis anterior**

| Transcript | Isoform | FVB_1 | FVB_2 | FVB_3 | FVB_4 | FVB_5 | A17_1 | A17_2 | A17_3 | A17_4 | A17_5 |
|---|---|---|---|---|---|---|---|---|---|---|---|
| ENSMUST00000022808 | Iso-201 | 31.03 | 18.70 | 28.09 | 32.55 | 25.88 | 9.99 | 0.00 | 12.37 | 14.89 | 14.41 |
| ENSMUST00000172557 | Iso-208 | 34.75 | 23.04 | 29.19 | 34.23 | 23.16 | 13.58 | 0.00 | 10.49 | 18.69 | 11.31 |
| ENSMUST00000116476 | Iso-202 | 9.90 | 18.48 | 9.22 | 7.19 | 11.92 | 28.76 | 23.21 | 18.27 | 13.30 | 19.96 |
| ENSMUST00000150975 | Iso-207 | 4.95 | 11.95 | 8.06 | 4.49 | 10.21 | 20.37 | 19.34 | 14.21 | 10.45 | 13.65 |
| ENSMUST00000172695 | Iso-209 | 19.88 | 28.27 | 33.48 | 24.38 | 21.28 | 18.97 | 14.89 | 12.97 | 16.03 | 15.59 |

**Muscle cell culture**

| Transcript | Isoform | WTA_1 | WTA_2 | WTA_3 | D7E_1 | D7E_2 | D7E_3 |
|---|---|---|---|---|---|---|---|
| ENSMUST00000022808 | Iso-201 | 47.84 | 27.89 | 39.15 | 95.27 | 59.93 | 32.02 |
| ENSMUST00000172557 | Iso-208 | 60.77 | 32.16 | 30.19 | 113.91 | 65.50 | 42.11 |
| ENSMUST00000116476 | Iso-202 | 34.48 | 14.45 | 48.62 | 27.16 | 33.41 | 47.06 |
| ENSMUST00000150975 | Iso-207 | 25.08 | 14.45 | 24.31 | 18.10 | 27.84 | 43.70 |

| | | | | | | | |
|---|---|---|---|---|---|---|---|
| ENSMUST00000172695 | Iso-209 | 37.61 | 61.40 | 48.62 | 27.16 | 83.52 | 43.70 |

**Table S6: List of OPMD and A17.1 overlapping muscle genes with APA-shift**

| Shift to proximal | Shift to distal |
|---|---|
| CSDE1 | MBNL1 |
| TMA7 | MEF2C |
| CALM1 | |
| HSPA9 | |
| NEB | |
| VCP | |
| TPM2 | |
| TNNT3 | |
| GAPDH | |

### B. Supplementary figures

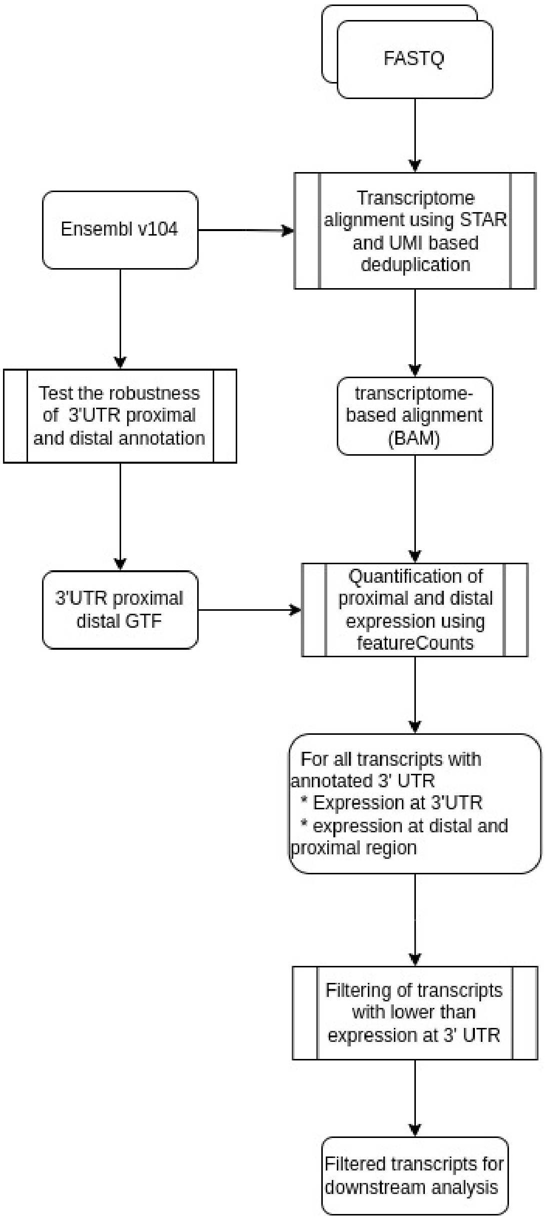


**Figure S1: A flow-chat summary of the bioinformatic steps that were used for the generation of reads for APA-shift calculation.**

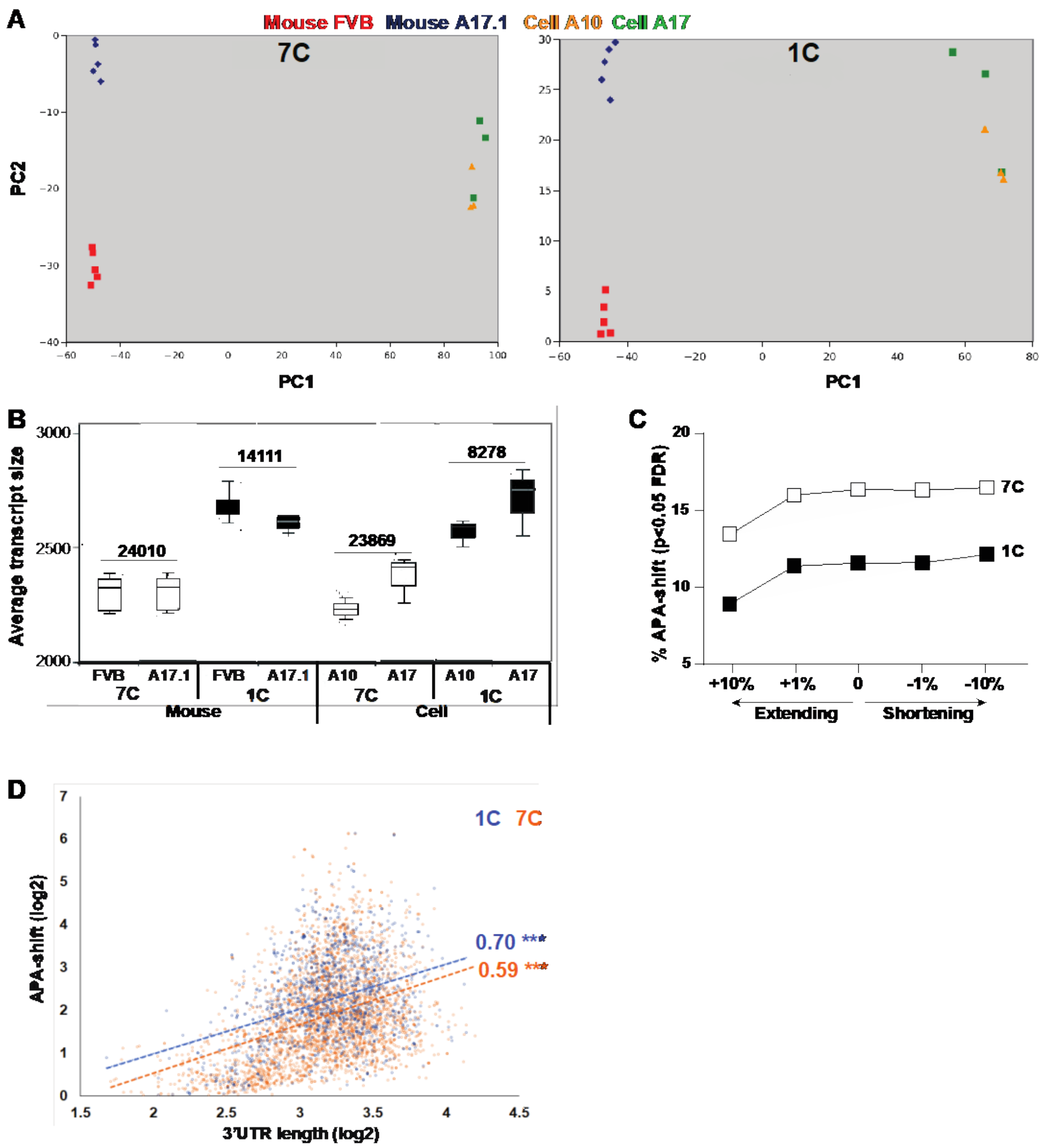


**Figure S2. 1C vs. 7C library prep results in mouse and cell OPMD models.**

- **A.** PCA plots of 7C or 1C RNAseq in mouse (FVB; A17.1) or mouse cells (A10; A17).
- **B.** Boxplot of 7C and 1C average insert size in mouse and cell models. The number of transcripts passing exclusion criteria is depicted.
- **C.** The percentage of transcripts with APA-shift (p<0.05, FDR) in mouse samples using different separation proximal and distal regions at the 3'UTR. 7C is denoted with open squares and 1C with close squares.
- **D.** Dot plot of 3'UTR length vs APA-shift (log2) of transcripts with APA-shift<0.05, FDR, in 7C and 1C datasets. The linear regression line is depicted with a dashed line, and the slope and significance (*** p<0.0001) are shown next to the slople.

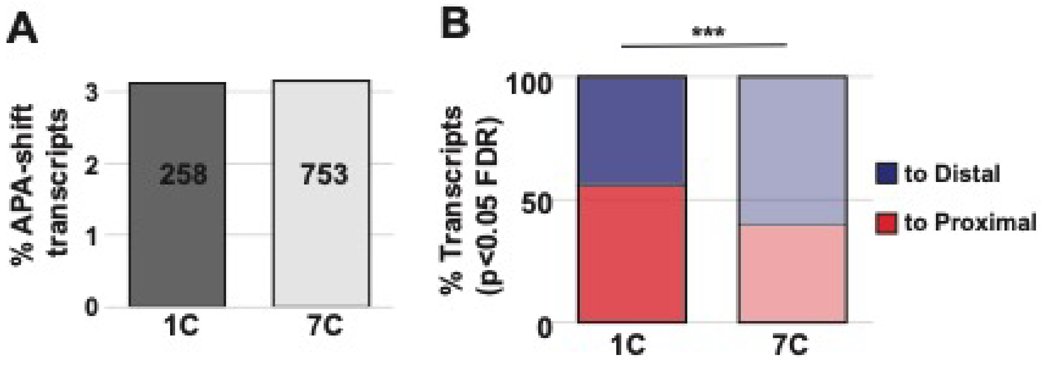


**Figure S3. 1C vs. 7C APA-seq in an OPMD muscle cell model**

- **A.** Bars show the percentage of transcripts with APA-shift from total transcripts in 1C or 7C datasets.
- **B.** Bars show percentage of transcripts proximal (red) or distal (blow) APA-shift from the significant (p<0.05, FDR) APA-shift transcripts in 1C or 7C datasets. A statistical difference between 1C and 7C shift direction was assessed with the chi-square test; *** p<0.0001.

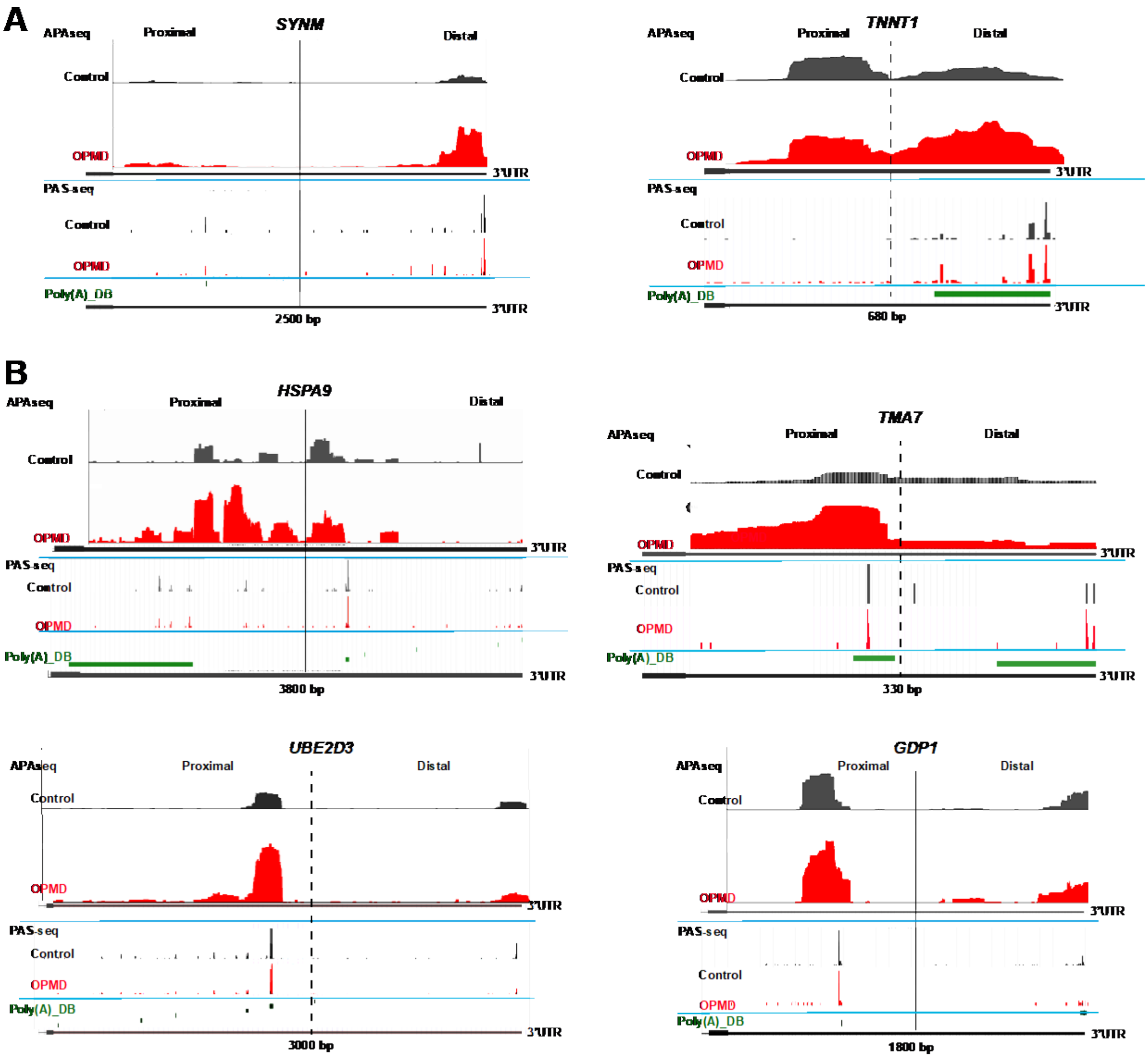


**Figure S4: IGV and PAS-seq comparison in six transcripts**

Transcripts examples are for shift to distal (A) or shift to proximal (B). Fer transcript, IGV in control (black) and OPMD (red) are on top followed by PAS-seq and PAS from poly(A)_DB (in green). The ensemble annotation 3'UTR is at the bottom, the length of the 3'UTR is depicted and a dashed line separates the proximal from distal regions.

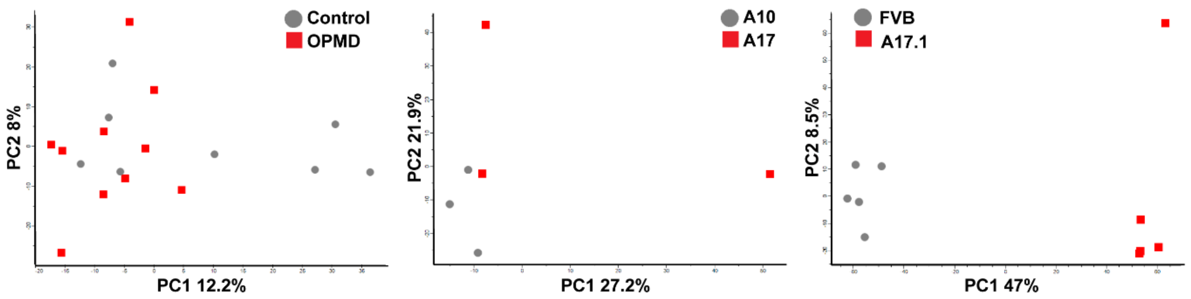


**Figure S5. PCA plots of APA-shift values in 1C samples, from left to right: human (controls and OPMD), muscle cell models (A10 and A17) , and mouse model (FVB and A17.1).** The percentage of variance for the first two principal components (PC) is depicted.
